# The Burden of Hypertension (HTN) and Diabetes Mellitus Among People Living with HIV and the General Population in Two Urban Cities in Zimbabwe: A Cross-Sectional Analysis of Integrated Screening Program Data (December 2022–December 2024)

**DOI:** 10.1101/2025.10.07.25337477

**Authors:** MacDonald Hove, Kudakwashe Collin Takarinda, Pauline Chimungu, Tinashe Theonevus Chinyanga, Pugie Tawanda Chimberengwa

## Abstract

**Introduction:** A pilot project was implemented between December 2022 and December 2024, to integrate HTN and DM screening into Zimbabwe’s clinical HIV program in Bulawayo and Chitungwiza. This sought to screen and determine the burden of non-communicable diseases (NCDs) among people living with HIV (PLHIV) as well as the general population aged ≥40 years. We set out to assess and compare HTN and DM prevalence among PLHIV and the general population (aged ≥40) in Zimbabwe.

**Methods:** A cross-sectional secondary data analysis utilizing routine program data was done. HTN and DM screening were integrated into HIV care services in public health facilities and in the community including outreach programs. Data on demographics, NCD risk factors, and screening outcomes were collected using the District Health Information Software (DHIS2) Tracker. All data were anonymized, securely stored, in compliance with ethical and data protection regulations.

**Results:** A total of 113,680 individuals were screened from December 2022 to December 2024, with 58% from the general population and 42% being PLHIV. Mean age was 53.1 (Q_1_=44.5: Q_3_=54.5) years. The prevalence of HTN among PLHIV was 48.2% compared to 46.4% in the general population; 47.6% among women compared to 46.3% in men (p<0.01). Overall prevalence of DM was 0.72% (821), with PLHIV having a prevalence of 0.44%, (209/47,624) compared to General population 0.93% (612/66,056), (p<0.05).

**Conclusion:** Majority of individuals screened for NCDs were PLHIV on ART, who also had a higher prevalence of HTN. The integration of NCDs screening and management with HIV programs is key for holistic patient centred care in chronic disease management. This has potential in improving outcomes for PLHIV while strengthening systems for general population. Collaborative efforts between government and partners are needed to equip primary care centres with resources (trained staff and equipment) to screen for NCDs, scale up integrated chronic disease management.

## BACKGROUND

Hypertension (HTN), commonly known as high blood pressure, is a significant global public health concern due to its role as a leading risk factor for cardiovascular diseases, stroke, and premature mortality.(1) Globally, an estimated 1.28 billion adults aged 30–79 years are affected by HTN, with approximately two-thirds residing in low- and middle-income countries. (2) About 21% of individuals with HTN have their condition under control, highlighting substantial gaps in awareness, treatment, and management.(2) In Africa, the burden of HTN is particularly pronounced. The prevalence among adults is estimated to be around 30.8%, with figures in Sub-Saharan Africa ranging between 30.0% and 31.1%.(3) Notably, certain population groups exhibit even higher rates; for instance, Central Africa reports a prevalence of 77.6% among individuals with DM.(4) Factors associated with this increased prevalence include urbanization, lifestyle changes, and limited access to healthcare services.(5) The HTN control rates in Africa are sub-optimal, with estimates suggesting that only 7.3% of individuals have their blood pressure adequately managed.(5)

Globally, Human Immunodeficiency Virus (HIV) remains a significant public health issue, with an estimated 38 million people living with HIV (PLHIV) in 2021.(6) In Sub-Saharan Africa, where approximately 70% of the world’s PLHIV reside, the prevalence of HIV is estimated at 10.5% among individuals aged 15–49 years.(6) Zimbabwe has achieved HIV epidemic control among adults, with 95% of PLHIV aware of their status, 99% of those diagnosed on ART, and 96% of those on ART achieving viral suppression.(6) As Anti-retroviral therapy (ART) coverage increases, a growing proportion of PLHIV are aging, with approximately half of ART clients now aged 40 years or older. This demographic shift raises the risk of non-communicable diseases (NCDs), as modelling studies estimated that the proportion of PLHIV with at least one key NCD would increase from 33% in 2015 to 47% by 2035. In comparison, among HIV-negative individuals, the prevalence was projected to rise from 14% to 26% over the same period.(7,8)

A systematic review and meta-analysis conducted in sub-Saharan Africa reported that among PLHIV, the most prevalent non-communicable diseases were obesity (32.2%), depression (30.4%), hypertension (20.1%), and hypercholesterolemia (21.3%).(9) In Zimbabwe, three studies based on routine health facility data have revealed a high burden of HTN, with prevalence rates ranging from 19.5% among individuals aged ≥18 years to 41% in those aged ≥50 years, while the prevalence of DM ranged from 1.4% to 8.4% in individuals aged ≥16 and ≥18 years, respectively.(10,11) Beyond the HIV-positive population, NCDs are a growing public health concern in Zimbabwe and across Sub-Saharan Africa. Thus, HTN and DM are among the leading NCDs affecting the general population, driven by aging, urbanization, and lifestyle changes. Both conditions contribute to a rising burden of cardiovascular diseases, disability, and mortality. Strengthening routine NCD screening and prevention strategies is critical to addressing health challenges in both HIV-positive and HIV-negative populations.

Type 2 Diabetes Mellitus (T2DM) has emerged as a significant public health challenge worldwide, with escalating prevalence rates that underscore the need for comprehensive prevention and management strategies. As of 2024, the global burden of DM has reached unprecedented levels. Recent data indicate that over 800 million adults worldwide are living with DM, marking a more than fourfold increase since 1990. This surge reflects a substantial rise in age-standardized prevalence across numerous countries, with notable increases observed in both men and women (9–11). Projections indicate that the number of DM cases could reach 1.3 billion by 2050, emphasizing the urgent need for effective interventions(10).

In Zimbabwe, DM prevalence has risen significantly over the past four decades (9,11). According to the 2024 estimates from the International Diabetes Federation (IDF), Zimbabwe currently has an adult (20–79 years) DM prevalence of 1.5%, affecting approximately 106,500 individuals (15). However, some national studies suggest higher prevalence rates, with a systematic review reporting 5.7% in certain populations (7). These discrepancies likely reflect differences in study methodologies, diagnostic criteria (e.g., HbA1c vs. fasting glucose), and regional variability in risk factors such as urbanization and obesity trends. Notably, undiagnosed DM remains a major concern, with an estimated 68.8% of cases undetected in Zimbabwe (15). Projections indicate that DM cases could rise to 239,300 by 2050 if current trends persist, underscoring the need for improved screening and preventive strategies (15).

The Organization for Public Health Interventions and Development (OPHID), with catalytic funding from the SANOFI Global Health Program, launched an HIV/NCD integration project focused on screening for HTN and DM among PLHIV in ART care and the at-risk general population aged ≥40 years. We hypothesized that PLHIV would have higher HTN prevalence while DM prevalence would be lower than the general population. This paper describes the demographic characteristics of those screened, monthly screening trends, prevalence of HTN and DM, and the associated factors among individuals screened between December 2022 and December 2024.

## METHODS

### Study Design

We conducted a cross-sectional secondary analysis using routinely collected program data.

### Setting

This project was part of OPHID-Sanofi’s technical and financial support to the Ministry of Health and Child Care (MOHCC), in Zimbabwe. This project was conducted in two urban areas of Zimbabwe, Bulawayo and Chitungwiza, where OPHID was implementing the United States President’s Emergency Plan for AIDS Relief (PEPFAR)-funded Target-Accelerate and Sustain Quality HIV Care for HIV epidemic control (TASQC) program. Bulawayo, the second-largest city in Zimbabwe, has a population of around 666,000, with 18% aged ≥40 years old,(16) and is served by 25 public health facilities providing ART care to 74,188 PLHIV by December 2024. These facilities offer integrated general health and HIV care services. Chitungwiza, a high-density dormitory town near Harare, has a population of approximately 371,000, with 18% aged ≥40 years old.(15) It hosts five public health facilities and one private hospital, providing ART services to 35,080 PLHIV by December 2024.

### Description of NCD integration into HIV program

This study was based on a cross-sectional secondary analysis of data collected from the implementation of an integrated HTN and DM screening program within HIV services in Zimbabwe. The program was rolled out between December 2022 and December 2024, aiming to strengthen the early detection and management of NCDs in urban settings.

#### Capacity Building

Community health workers (CHWs) and facility-based nurses were trained to measure blood pressure and blood sugar as part of routine screening activities in both health facility and community outreach settings. Blood pressure and blood glucose measurements were done using automated, home-use digital devices. Blood pressure was measured using validated, automated digital sphygmomanometers (Omron® model), following standard protocols. Measurements were taken in a seated position after at least five minutes of rest. In both community settings and facility settings, two readings were recorded and averaged where possible, otherwise the lower reading was used.

Capillary blood glucose levels were measured using portable glucometers, using finger-prick samples. A random blood glucose level ≥11.1 mmol/L or fasting blood glucose ≥7.0 mmol/L was used to define suspected DM, in line with the WHO criteria. All devices used were battery-operated and designed for home or field use, allowing for mobility during community-based outreach. Prior to implementation, CHWs were equipped with these devices and trained in standardized procedures for accurate measurement, calibration, infection prevention, and data recording.

Additionally, it was observed that nurses at the primary healthcare facilities were not confident to manage HTN and DM, with only doctors being authorized to initiate hypertensive and diabetic medicines at law. To address these gaps, a structured mentorship and training program was introduced. Health facility staff, including nurses, received targeted training on the management of HTN and DM. The program procured BP machines, glucometers, and batteries for health facilities to ensure availability of screening equipment. Locum arrangements were made to guarantee the availability of doctors and nurses and ensure seamless service delivery for recipients of care (RoC).

#### Addressing Treatment Gaps and Supply Chain Management

While the screening and linkage to treatment improved, a new challenge emerged, clients diagnosed with HTN and DM faced difficulties in accessing medicines at public health facilities due to long-term stockouts. Medicines were often unavailable due to the underestimation of NCD medicine needs during national medicines quantification exercises, compounded by poor data quality in reporting systems. To mitigate this, technical support was provided for the national medicine quantification process, ensuring a more accurate and data driven estimation of medicine needs was provided during the process.

Additionally, to bridge immediate gaps in medication access, the program facilitated linkages between public health facilities and private pharmacies, negotiating discounted prices for recipients of care (RoCs). In some cases, donations of essential NCD medications were secured from wholesale pharmacies to supplement public supply shortages. Pooled procurement of medicines for RoCs was enabled for cheaper medicines from pharmacies who supplied cheaper “access medicines” procured through the Sanofi Global Health Unit.

### Data variables and sources of data

Routine program data collected included the date of screening, the cadre who conducted the screening (e.g., CHW or nurse), gender, district, and the type of patient (general population or PLHIV). Data on NCD risk factors were also recorded, including: i) engagement in physical activity for ≥30 minutes daily, ii) experiencing polyuria, iii) experiencing polydipsia, iv) tobacco use, and v) alcohol use. Blood pressure readings (systolic and diastolic in mmHg) and blood glucose levels (mmol/L) were also measured. All data were entered into a customized DHIS2 Tracker Capture application, which was installed on electronic tablets and mobile phones.

A comprehensive data reporting system was developed to improve the quality of NCD data. A new MOHCC Monthly Reporting Form (MRF) was introduced, incorporating disaggregated NCD indicators by age and sex. This allowed for more precise tracking of HTN and DM trends for PLHIV. These tools were adopted by the MOHCC for use by all health facilities in the country.

### Data analysis

Data were downloaded from a DHIS2 server and cleaned in Microsoft Excel before being imported into Stata (Version 17.0) for statistical analysis. The data was downloaded on 20 March 2025. The primary outcomes, ‘being hypertensive’ and ‘being diabetic,’ were defined as follows: HTN was having a systolic blood pressure >140 mmHg and/or a diastolic blood pressure >90 mmHg, while DM was defined as a random blood sugar level >11.1 mmol/L. The prevalence of HTN and DM was determined by dividing the number of hypertensive and prediabetic individuals (from December 2022 to December 2024) by the total number of individuals screened for NCDs during the same period.

Descriptive statistics were used to summarize participant characteristics, including age, sex, HIV status, and screening outcomes. Categorical variables, such as HTN and DM status, sex, and HIV status, were expressed as frequencies and percentages. Continuous variables, such as age, were summarized using means, ranges and standard deviations. Comparative analyses were conducted to assess differences in the prevalence of HTN and DM between subgroups. Chi-square (χ²) tests were used to assess associations between categorical variables, including the comparison of HTN and DM prevalence by HIV status and sex. Statistical significance was set at a two-sided p-value of <0.05. In addition, trends in NCD prevalence by age group were analysed to identify potential age-related patterns in disease burden. All statistical analyses were conducted using Microsoft Excel and Stata version 17 (StataCorp, College Station, TX, USA). Data were de-identified prior to analysis to ensure confidentiality. Only authorized personnel had access to the data, and all analyses were performed in compliance with ethical standards and data protection protocols. A significance level of 5% was applied.

### Ethics approval

The organisation was given an ethical approval by the Ministry of Health and Child Care to implement and collect data for the project. Data safety and confidentiality were prioritized throughout the study. All participant information was anonymized and securely stored in compliance with ethical guidelines and data protection regulations. Only authorized personnel had access to the data, which was stored on password-protected electronic platforms. Any identifiable client information was removed before analysis, ensuring that privacy was maintained at all stages of the study. The authors did not have access to data with personal identifiable information. The study adhered to ethical standards set by the Ministry of Health and Child Care, Zimbabwe.

## RESULTS

A total of 113,680 clients over the age of 40 years were screened between December 2022 and December 2024. Females constituted 62.42% and 37.58% were males. The mean age of participants was approximately 53.1 years. Participants ranged in age from 40 to 95 years (Q_1_=44.5: Q_3_=54.5). The largest proportion of individuals screened fell within the 40–49-year age group, accounting for 45.7% of the sample. This was followed by 28.6% in the 50–59-year group, and 16.5% in the 60–69-year group. Geographically, most of the screened clients were from Bulawayo, 63.48%, with the remaining 36.52% from Chitungwiza. The general population accounted for 58% of the sample, while PLHIV on ART made up 42% of those screened. The demographic results are summarized in Table 1.

**Table 1:** Demographic characteristics among clients over 40 years screened for HTN and DM in Bulawayo and Chitungwiza, December 2022 - December 2024.

| Variable | n= | (%) |
| --- | --- | --- |
| **Total** | 113,680 | (100) |
| **Sex** |  |  |
| Female | 70,956 | (62.42) |
| Male | 42,724 | (37.58) |
| **Age group (in years)** |  |  |
| 40-49 | 51,894 | (45.65) |
| 50-59 | 32,494 | (28.58) |
| 60-69 | 18,764 | (16.51) |
| 70-79 | 6,104 | (5.37) |
| 80-89 | 1,484 | (1.31) |
| >=90 | 182 | (0.16) |
| Not recorded | 2,758 | (2.43) |
| **District** |  |  |
| Bulawayo | 72,165 | (63.48) |
| Chitungwiza | 41,515 | (36.52) |
| **Type of client** |  |  |
| General population | 66,056 | (58) |
| PLHIV on ART | 47,624 | (42) |

ART = antiretroviral therapy; PLHIV = people living with Human Immunodeficiency Virus

Table 2 shows the prevalence of NCD risk factors among clients screened for HTN and DM in Bulawayo and Chitungwiza from December 2022 to December 2024. Most clients, 76.83%, reported engaging in physical activity for at least 30 minutes daily, while 22.97% did not. Regarding symptoms of DM, 89.55% of participants did not report polyuria (frequent urination), and 90.14% did not experience polydipsia (excessive thirst), with 6.19%, and 5.38% reporting these symptoms, respectively.

**Table 2:**
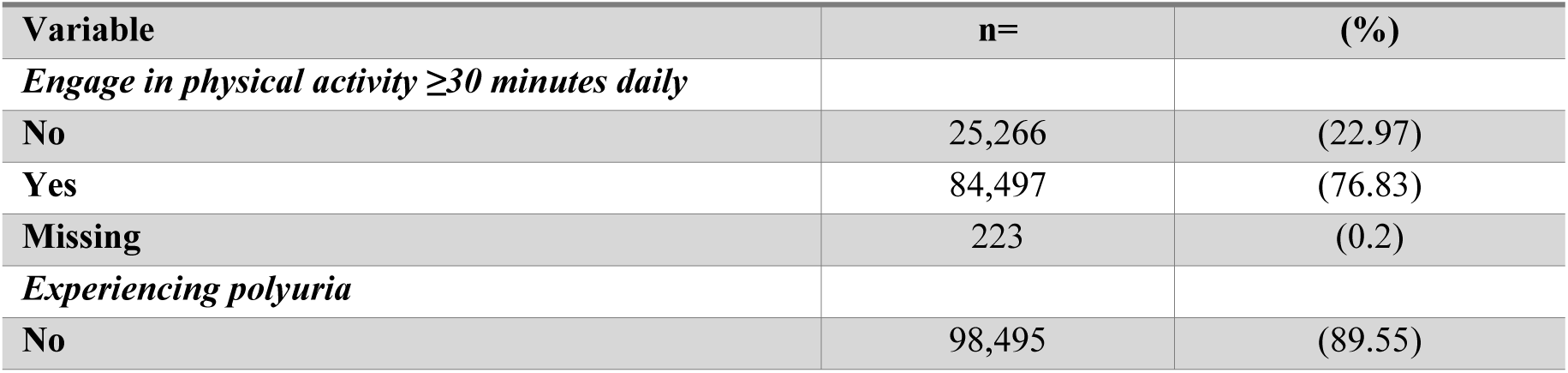
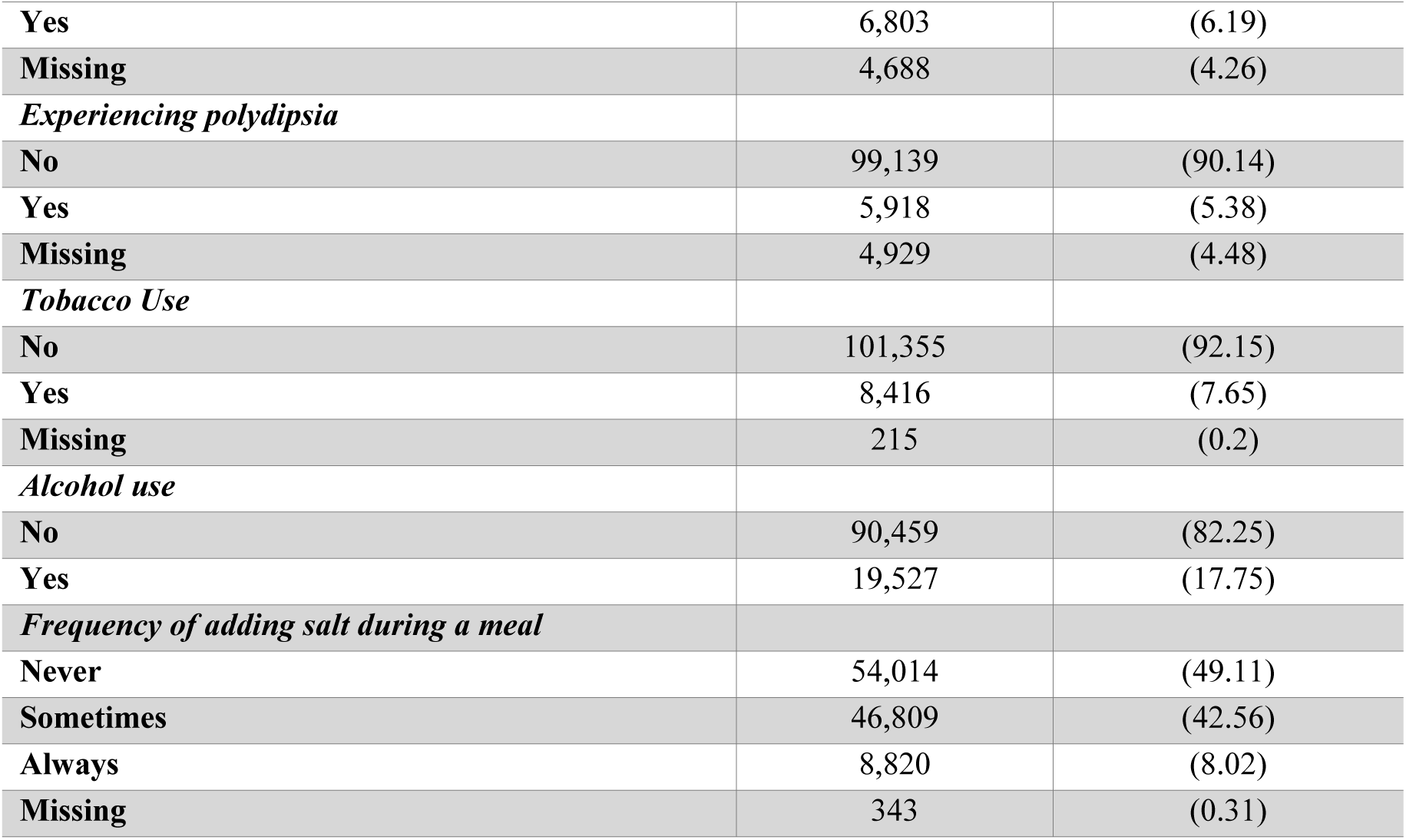
Prevalence of NCD risk factors among clients over 40 years screened for HTN and DM in Bulawayo and Chitungwiza from December 2022 to December 2024.

| Variable | n= | (%) |
| --- | --- | --- |
| **Engage in physical activity $\geq 30$ minutes daily** |  |  |
| No | 25,266 | (22.97) |
| Yes | 84,497 | (76.83) |
| Missing | 223 | (0.2) |
| **Experiencing polyuria** |  |  |
| No | 98,495 | (89.55) |
| **Yes** | 6,803 | (6.19) |
| **Missing** | 4,688 | (4.26) |
| *Experiencing polydipsia* |  |  |
| **No** | 99,139 | (90.14) |
| **Yes** | 5,918 | (5.38) |
| **Missing** | 4,929 | (4.48) |
| *Tobacco Use* |  |  |
| **No** | 101,355 | (92.15) |
| **Yes** | 8,416 | (7.65) |
| **Missing** | 215 | (0.2) |
| *Alcohol use* |  |  |
| **No** | 90,459 | (82.25) |
| **Yes** | 19,527 | (17.75) |
| *Frequency of adding salt during a meal* |  |  |
| **Never** | 54,014 | (49.11) |
| **Sometimes** | 46,809 | (42.56) |
| **Always** | 8,820 | (8.02) |
| **Missing** | 343 | (0.31) |

Tobacco use was reported by 7.65% of clients, with 92.15% were reportedly non-smokers while alcohol consumption was reported by 17.75% of clients. Dietary habits related to salt consumption showed that 49.11% of clients never added salt to their meals, 42.56% sometimes added salt, and 8.02% always added salt.

Overall, as shown in Table 3 HTN was present in 47.1% of the screened population, with a higher prevalence among HIV-positive individuals (48.2%) compared to HIV-negative individuals (46.4%), a difference that was statistically significant (p<0.05). DM was less common overall (0.72%); however, DM prevalence was significantly lower in HIV-positive individuals (0.44%) as compared to HIV-negative individuals (0.93%), (p<0.05). The combined prevalence of both HTN and DM was reported at 47.3%, with a higher proportion in HIV-positive individuals (48.3%) compared to HIV-negative individuals (46.7%), and this difference was statistically significant (p<0.05). The results indicate variations in NCD prevalence based on HIV status, with HTN being more common among HIV-positive individuals and DM being less common in this group, while the co-morbidity of both conditions followed a similar trend to HTN alone.

**Table 3:** Comparison of NCD prevalence between HIV positive and negative people above 40 years screened for NCDs in Bulawayo and Chitungwiza, December 2022-December 2024.

| Outcomes | Overall | HIV positive | HIV negative | p-value |
| --- | --- | --- | --- | --- |
|  | n/N (%) | n/N (%) | n/N (%) |  |
| **HTN** | 53,622/113 680<br>(47.1%) | 22,954/47,624 (48.2%) | 30,668/66,056 (46.5%) | <0.05 |
| **DM** | 821/113 680 (0.72%) | 209/47624 (0.44%) | 612/66 056 (0.93%) | <0.05 |
| **HTN & DM** | 53,880/113,680<br>(47.3%) | 23,010/47,624 (48.3%) | 30,870/66,056 (46.7%) | <0.05 |

## DISCUSSION

The results from this screening highlight the substantial burden and prevalence of NCDs, particularly HTN and DM, among urban populations in Zimbabwe. This discussion focuses on interpreting these findings, with particular attention to differences observed between PLHIV and the general population, and the broader implications for public health.

A key finding from the analysis of this program data is that 42% of individuals screened for HTN and DMs were PLHIV who were on ART. This high proportion reflects the successful integration of NCD screening into HIV care program. Given the increased risk of NCDs among PLHIV—driven by factors such as chronic inflammation, metabolic side effects of long-term ART, and aging—routine screening for hypertension and diabetes in this population is essential (17,18). Bulawayo and Chitungwiza have effectively adopted a differentiated, person-centered approach that extends care beyond viral suppression to broader health outcomes, aligning with WHO recommendations on integrated chronic disease management.

One important factor contributing to the high proportion of PLHIV among individuals screened for hypertension and diabetes in this study is their frequent and structured contact with health services. Routine HIV care involves regular clinical visits for ART refills, viral load monitoring, and opportunistic infection screening, providing repeated opportunities for additional health interventions, including NCD screening (19). This ongoing engagement facilitates early detection and management of co-morbid conditions, which is especially critical as PLHIV are at increased risk of developing NCDs due to aging, ART side effects, and chronic immune activation (2). As a result, PLHIV have better access to integrated care pathways and have a higher likelihood of being screened for NCDs compared to individuals in the general population, who may only engage with the health system when symptomatic or acutely ill. While this reflects effective leveraging of HIV care platforms for broader health service delivery, it also underscores potential inequities in access to preventive services. Individuals not living with HIV may be missed by vertical health service delivery models, emphasizing the need to strengthen community outreach and integrated NCD screening into primary health care to ensure more equitable coverage. Ultimately, the frequent health system contacts that PLHIV experience serves as a platform for improved chronic disease care management, a model for delivering comprehensive, person-centred care.

In this NCD screening program, men constituted 37.6% of individuals who were screened for HTN and DM in Bulawayo and Chitungwiza between December 2022 and December 2024. Although this figure is markedly lower than that of women (62.4%), it represents a relatively higher male participation rate compared to similar NCD screening initiatives in sub-Saharan Africa, where male representation typically ranges between 20% and 35% (12,20). The modestly improved male uptake observed in this program reflect effective outreach strategies, workplace wellness initiatives, and integration of screening into routine services accessed by both sexes.

Globally and regionally, men often face multiple barriers to accessing preventive health services, including limited health awareness, cultural norms that discourage health-seeking behavior, and occupational commitments that restrict time availability (18). As a result, NCD screening programs may inadvertently underserve men if these barriers are not addressed. To build on the relatively higher male participation in this study, future programming should consider leveraging male-dominated spaces such as workplaces, markets, and social clubs to offer screening. Additionally, targeted health communication and the use of male peer educators could further increase male engagement in NCD prevention services. This approach aligns with growing recognition that equitable access to preventive care is essential to controlling the growing burden of hypertension and diabetes in low and middle-income countries.

The high prevalence of HTN observed in this study, 47.1% overall among individuals aged 40 to 95 years, aligns with regional trends indicating increased cardiovascular risk in ageing populations, particularly in sub-Saharan Africa. The slightly higher prevalence among HIV-positive individuals (48.2%) compared to HIV-negative individuals (46.4%), suggests a potential association between HIV infection or its long-term treatment and elevated blood pressure. Several studies have demonstrated that PLHIV are at increased risk of hypertension, likely due to chronic immune activation, endothelial dysfunction, and the metabolic side effects of ART (21). Moreover, with improved ART access and survival, PLHIV are living longer, which naturally increases their risk of age-related conditions such as hypertension.

These findings underscore the urgent need for integrating hypertension screening and management into HIV care services. Given the high burden and shared risk factors such as obesity, sedentary lifestyles, and smoking, there is a practical and cost-effective rationale for a “one-stop-shop” approach to chronic disease care. Previous implementation experiences in sub-Saharan Africa have shown that such integration can improve early detection and clinical outcomes for both communicable and non-communicable diseases (12). Additionally, the high overall prevalence highlights the importance of public health interventions aimed at primary prevention, community awareness, and lifestyle modification strategies targeting the general population.

As for DM, while less prevalent overall than hypertension in this cohort, it showed a noteworthy and statistically significant difference between HIV-positive (0.44%) and HIV-negative individuals (0.93%). Interestingly, the higher prevalence among HIV-negative individuals may suggest that HIV infection or ART might not be as strongly associated with diabetes as previously hypothesized. Prior studies have yielded mixed findings: some report higher rates of DM among PLHIV, potentially due to protease inhibitors and lipodystrophy (22), while others have found no significant difference when controlling for age and BMI (23).

Despite the lower observed prevalence, the presence of DM in 821 individuals underscores its growing burden in Zimbabwe’s ageing population. The dual burden of communicable and non-communicable diseases presents a significant challenge to a health system that historically prioritized communicable disease management. As such, expanding NCD screening, particularly for DM, into existing HIV platforms and general primary care services is imperative. Ensuring the availability of affordable glucose testing, follow-up care, and education about modifiable risk factors is critical to preventing complications such as neuropathy, nephropathy, and cardiovascular disease (18). Additionally, further research is needed to explore barriers to DM detection and care among PLHIV, which may influence the current prevalence figures.

The study’s reliance on random blood glucose (RBG) measurements alone likely led to a significant underestimation of the true burden of DM in the population. While an RBG ≥11.1 mmol/L is a WHO-recognized diagnostic threshold, it has substantially lower sensitivity compared to fasting blood glucose (FBG), oral glucose tolerance testing (OGTT), or glycated haemoglobin (HbA1c), which are considered gold-standard diagnostic methods (22). Reference to RBG may miss individuals with isolated fasting hyperglycaemia or impaired glucose tolerance, conditions that reflect early or subclinical disease stages (12). Consequently, the observed DM prevalence of 0.72% is markedly lower than national estimates for Zimbabwe, which range from 1.5% to 5.7%. This may be indicating the likelihood of systematic underdiagnosis and underreporting of disease burden. However, the study population was not representative of national sample dynamics and thus this may still be a true representation in this population. Studies in similar low-resource settings suggest that RBG-based screening captures only 30% to 50% of cases that OGTT or HbA1c would detect (22). Moreover, among high-risk groups such as PLHIV, chronic inflammation and ART can further distort glucose metabolism, complicating disease detection and early diagnosis. A more accurate assessment of DM burden, particularly in high-risk populations, would require a multi-test diagnostic algorithm incorporating FBG and HbA1c.

## CONCLUSION

The integration of NCD screening into existing HIV and primary care in Bulawayo and Chitungwiza effectively identified a high burden of hypertension, particularly among PLHIV. By leveraging the structured, regular contact PLHIV have with health services for ART refills and clinical monitoring, the program expanded its scope to holistic, person-centred chronic disease management. This approach demonstrates the value of collaborative, cross-program strategies in tackling the growing NCD epidemic in Zimbabwe, aligning with WHO’s recommendations for integrated care. However, HIV-negative individuals remain relatively underserved, underscoring the need to expand screening into primary health care and workplace wellness programs. While HTN emerged as the most prevalent NCD in the screened population, DM was detected at a much lower rate than expected, likely due to reliance on random blood glucose testing alone. This finding highlights the need for improved screening efforts that use more sensitive diagnostic tools such as fasting blood glucose and HbA1c and targeting at risk populations within health facilities. Addressing this high burden of HTN requires collaborative efforts are between the Ministry of Health and partners to provide equipment for screening and capacity building at primary care level. A comprehensive, primary-care focused integrated, and person-centered model linking HIV, NCD, and community health offers the best pathway to addressing the dual burden of NCDs and HIV in Zimbabwe’s ageing and increasingly urban population.

## ACKNOWLEDGEMENTS

The authors wish to express gratitude to; Ministry of Health and Child Care Zimbabwe, Bulawayo City and Chitungwiza City councils which own and support the health facilities with human resources and medical products. The authors also appreciate PEPFAR through USAID for funding the TASQC program which supports the 2 districts where the screening was done, to the dedicated nurses, community health workers, and health information officers whose unwavering commitment to RoC and accurate reporting makes a profound impact on healthcare delivery. Their tireless efforts in screening, treatment, and data management were invaluable in ensuring high-quality care and evidence-based decision-making. This article is a testament to their hard work and dedication, and it would not have been possible without their contributions. Their role in strengthening health systems and improving patient outcomes, particularly in the integration of HIV and NCD care, is truly commendable.

## Data Availability Statement

Data cannot be shared publicly due to Zimbabwe Ministry of Health restrictions. De-identified data is available upon request to OPHID after signing a data access agreement.

